# Multi-ancestry polygenic risk scores for venous thromboembolism

**DOI:** 10.1101/2024.01.09.24300914

**Authors:** Yon Ho Jee, Florian Thibord, Alicia Dominguez, Corriene Sept, Kristin Boulier, Vidhya Venkateswaran, Yi Ding, Tess Cherlin, Shefali Setia Verma, Valeria Lo Faro, Traci M. Bartz, Anne Boland, Jennifer A. Brody, Jean-Francois Deleuze, Joseph Emmerich, Marine Germain, Andrew D. Johnson, Charles Kooperberg, Pierre-Emmanuel Morange, Nathan Pankratz, Bruce M. Psaty, Alexander P. Reiner, David M. Smadja, Colleen M. Sitlani, Pierre Suchon, Weihong Tang, David-Alexandre Trégouët, Sebastian Zöllner, Bogdan Pasaniuc, Scott M. Damrauer, Serena Sanna, Harold Snieder, Lifelines Cohort Study, Christopher Kabrhel, Nicholas L. Smith, Peter Kraft, INVENT Consortium

**Affiliations:** Department of Epidemiology, Harvard T.H. Chan School of Public Health, MA, USA; Population Sciences Branch, Division of Intramural Research, National Heart, Lung and Blood Institute, MD, USA; The Framingham Heart Study, 73 Mt. Wayte Ave, Suite #2, Framingham, MA, 01702 USA; Department of Biostatistics, University of Michigan, Ann Arbor, MI, USA; Department of Biostatistics, Harvard T.H. Chan School of Public Health, Boston, MA 02115, USA; Bioinformatics Interdepartmental Program, University of California Los Angeles, Los Angeles, CA, USA; Department of Oral Biology, University of California Los Angeles School of Dentistry, Los Angeles, CA, USA; Department of Pathology and Laboratory Medicine, University of Pennsylvania Perelman School of Medicine, Philadelphia, PA, USA; Department of Epidemiology, University of Groningen, University Medical Center Groningen, Groningen, The Netherlands; Department of Immunology, Genetics and Pathology, Science for Life Laboratory, Uppsala University, Uppsala, Sweden; Cardiovascular Health Research Unit, Departments of Biostatistics and Medicine, University of Washington, 4333 Brooklyn Ave, Seattle, WA 98195; Université Paris-Saclay, CEA, Centre National de Recherche en Génomique Humaine, Evry, France; Laboratory of Excellence in Medical Genomics, GENMED, Evry, France; Cardiovascular Health Research Unit, Department of Medicine, University of Washington, 4333 Brooklyn Ave, Seattle, WA 98195; Centre d’Etude du Polymorphisme Humain, Fondation Jean Dausset, Paris, France; Department of Vascular Medicine, Paris Saint-Joseph Hospital Group, University of Paris, Paris, France; UMR1153, INSERM CRESS, Paris, France; University of Bordeaux, INSERM, Bordeaux Population Health Research Center, UMR 1219, Bordeaux, France; Division of Public Health Sciences, Fred Hutchinbson Cancer Center, Seattle WA 98109; Aix-Marseille University, INSERM, INRAE, Centre de Recherche en CardioVasculaire et Nutrition, Laboratory of Haematology, CRB Assistance Publique – Hôpitaux de Marseille, HemoVasc, Marseille, France; Department of Laboratory Medicine and Pathology, University of Minnesota, Minneapolis, Minnesota, 55455, USA; Department of Epidemiology, University of Washington, 4333 Brooklyn Ave, Seattle, WA 98195; Department of Health Systems and Population Health, University of Washington, 4333 Brooklyn Ave, Seattle, WA 98195; Innovative Therapies in Hemostasis, Université de Paris, INSERM, F-75006 Paris, France; Hematology Department and Biosurgical Research Lab (Carpentier Foundation), Assistance Publique Hôpitaux de Paris, Centre-Université de Paris (APHP-CUP), F-75015 Paris, France; Division of Epidemiology and Community Health, School of Public Health, University of Minnesota, Minneapolis, Minnesota, 55454, USA; Department of Genetics, University of Pennsylvania Perelman School of Medicine, Philadelphia, PA, USA; Department of Surgery, Department of Genetics, and Cardiovascular Institute, Perelman School of Medicine, University of Pennsylvania, Philadelphia PA; Department of Surgery, Corporal Michael Crescenz VA Medical Center, Philadelphia PA; University of Groningen, UMCG, Department of Genetics, Groningen, the Netherlands; Institute for Genetics and Biomedical Research, National Research Council, Monserrato, Italy; Center for Vascular Emergencies, Department of Emergency Medicine, Massachusetts General Hospital, Harvard Medical School, Boston, MA, USA; Kaiser Permanente Washington Health Research Institute, Kaiser Permanente Washington, Seattle WA 98101, USA; Seattle Epidemiologic Research and Information Center, Department of Veterans Affairs Office of Research and Development, Seattle WA 98108, USA; Transdivisional Research Program, Division of Cancer Epidemiology and Genetics, National Cancer Institute, National Institutes of Health, MD, USA

## Abstract

Venous thromboembolism (VTE) is a significant contributor to morbidity and mortality, with large disparities in incidence rates between Black and White Americans. Polygenic risk scores (PRSs) limited to variants discovered in genome-wide association studies in European-ancestry samples can identify European-ancestry individuals at high risk of VTE. However, there is limited evidence on whether high-dimensional PRS constructed using more sophisticated methods and more diverse training data can enhance the predictive ability and their utility across diverse populations. We developed PRSs for VTE using summary statistics from the International Network against Venous Thrombosis (INVENT) consortium GWAS meta-analyses of European- (71,771 cases and 1,059,740 controls) and African-ancestry samples (7,482 cases and 129,975 controls). We used LDpred2 and PRSCSx to construct ancestry-specific and multi-ancestry PRSs and evaluated their performance in an independent European- (6,261 cases and 88,238 controls) and African-ancestry sample (1,385 cases and 12,569 controls). Multi-ancestry PRSs with weights tuned in European- and African-ancestry samples, respectively, outperformed ancestry-specific PRSs in European- (PRSCSX_EUR_: AUC=0.61 (0.60, 0.61), PRSCSX_combined_EUR_: AUC=0.61 (0.60, 0.62)) and African-ancestry test samples (PRSCSX_AFR_: AUC=0.58 (0.57, 0.6), PRSCSX_combined_AFR_: AUC=0.59 (0.57, 0.60)). The highest fifth percentile of the best-performing PRS was associated with 1.9-fold and 1.68-fold increased risk for VTE among European- and African-ancestry subjects, respectively, relative to those in the middle stratum. These findings suggest that the multi-ancestry PRS may be used to identify individuals at highest risk for VTE and provide guidance for the most effective treatment strategy across diverse populations.

## Introduction

Venous thromboembolism (VTE) is among the top five most common vascular diseases in most countries (1). The estimated lifetime risk of VTE is 8% among US adults (2). Approximately 20% of individuals die within 1 year of a VTE diagnosis often from the provoking conditions, and complications are common among survivors (3). Thus, the development of tools that stratify people according to their risk of developing VTE is helpful, which could inform risk-stratified prevention strategies that contribute to reducing the burden of VTE.

Polygenic risk scores (PRS) are useful tools for approximating the cumulative genetic susceptibility to complex traits and diseases. PRSs based on the independent genome-wide significant variants discovered in genome-wide association studies (GWAS) European-ancestry samples (4–9) have been demonstrated to identify individuals at high risk of VTE (10,11). However, there is limited evidence on whether high-dimensional PRS that are not restricted to genome-wide significant variants can enhance the predictive ability.

In the USA, the incidence of VTE is approximately 65% higher in those who identify as Black Americans than White Americans (12,13). Polygenic risk prediction models for VTE could be particularly important among Black Americans, as a clinical tool to reduce this disparity in VTE risk. (This does not preclude research into structural inequities and social determinants of health, which might inform policy interventions to reduce disparities between Black and White Americans.) However, previously developed VTE PRS have been optimized for European-ancestry populations, and their utility in other populations is unknown. In particular, we are unaware of any efforts to develop VTE PRS specifically for Black Americans.

We developed ancestry-specific and multi-ancestry PRSs for VTE leveraging large GWAS meta-analyses in European-and African-ancestry samples. We validated these PRSs by estimating relative VTE risks across PRS quintiles in five independent U.S.-based studies. We focus on PRS including common variants (minor allele frequencies above 1%) due to difficulties measuring or imputing low frequency or rare variants from GWAS data or imprecision of estimating rare variant associations. Thus our PRSs complement known low frequency variants (such as rs6205 in *F5*) or known clinical and behavioral risk factors. Here we concentrate on developing PRSs that perform well in diverse populations. Future work will be needed to (a) develop and evaluate models that combine these PRSs with low-frequency and rare variants and other risk factors and (b) assess the clinical utility of VTE risk models for targeted prevention, screening, or treatment (14,15).

## Results

### Study sample

The overall study design is illustrated in **Figure 1**. Our PRS development consisted of two steps: training ancestry-specific PRS and tuning multi-ancestry PRS. We trained ancestry-specific PRSs using European- and African ancestry GWAS summary statistics from the INVENT consortium and two Bayesian methods (LDPRED2(14) and PRSCSx(15)). We then tuned the constructed multi-ancestry PRSs by regressing VTE case-control status on a linear combination of the two ancestry-specific PRSs in two separate tuning samples: one European-ancestry tuning sample (1,329 cases and 1,324 controls) and one African-ancestry tuning sample (238 cases and 3,589 controls). The testing data set comprised 6,781 cases and 103,016 controls of European ancestry and 1,385 cases and 12,569 controls of African ancestry from five independent studies. Table S1 presents a brief summary of participating studies and biobanks, including basic information about each study or biobank (location, institute, cohort size, and sample recruiting approach), participants (ancestry and age), and genotypes (genotyping platforms and imputation reference).

**Figure 1.**
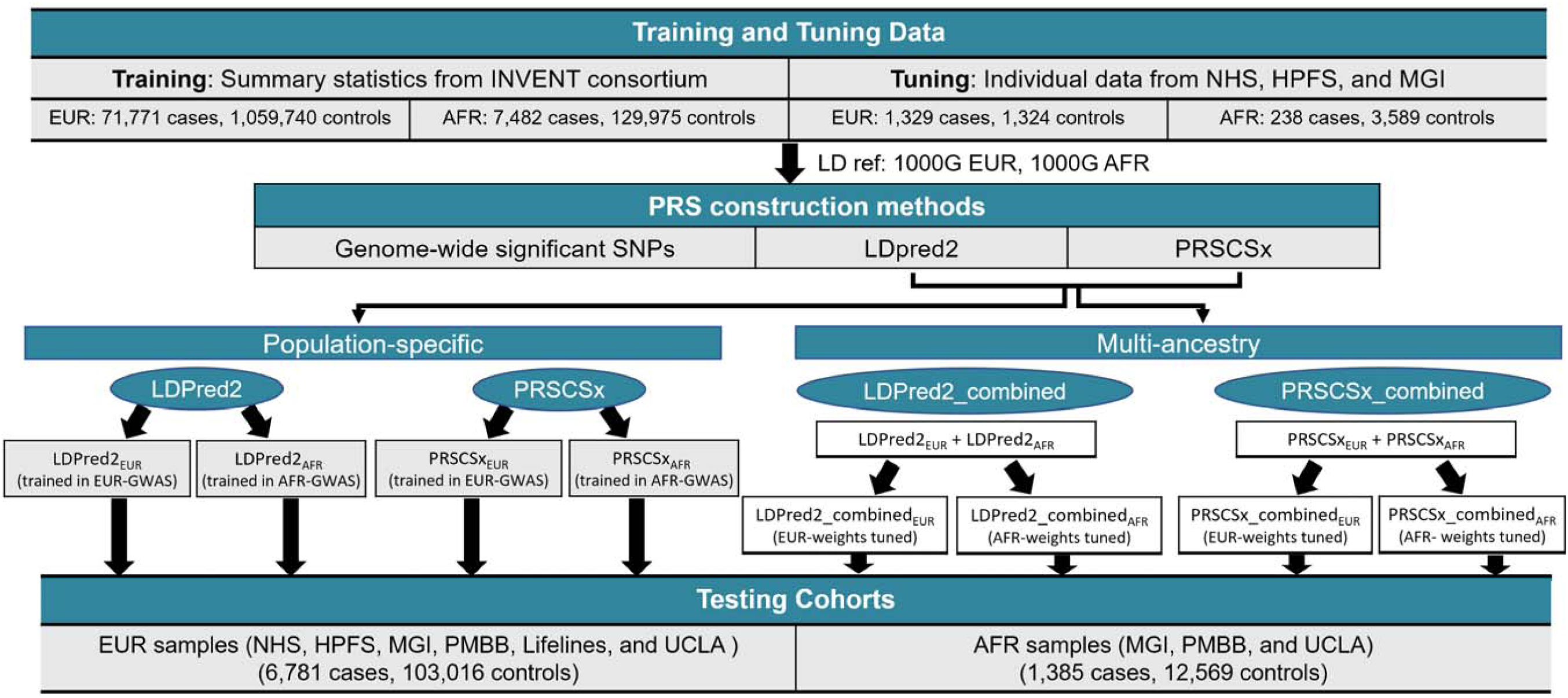
Overview of development and validation of population-specific and multi-ancestry PRS for venous thromboembolism.

### Comparing PRS distributions across populations

Four single-ancestry PRSs and four multi-ancestry PRSs for VTE were constructed using LDpred2 and PRSCSx and validated in independent European ancestry and African ancestry individuals: (i) LDpred2 trained using European-ancestry GWAS summary statistics (LDpred2_EUR_); (ii) LDpred2 trained using African-ancestry summary statistics (LDpred2_AFR_); (iii) PRS-CS trained using European-ancestry summary statistics (PRSCSX_EUR_); (iv) PRSCS trained using African ancestry summary statistics (PRSCSX_AFR_); and (v) LDpred2_EUR_ + LDpred2_AFR_ with weights tuned in an independent European-ancestry tuning sample; (vi) LDpred2_EUR_ + LDpred2_AFR_ with weights tuned in and independent African-ancestry tuning sample (LDpred2_combined_AFR_); (vii) PRSCSX_EUR_ + PRSCSX_AFR_ with weights tuned in the European-ancestry tuning sample (PRSCSX_combined_EUR_); (viii) PRSCSX_EUR_ + PRSCSX_AFR_ with weights tuned in the African-ancestry tuning sample (PRSCSX_combined_AFR_). All PRSs had higher means in cases than controls in the test data sets (**Table 1**). Among the European-ancestry VTE cases, the mean PRS was higher for the PRS tuned in European-ancestry samples than for the PRS tuned in African-ancestry samples. The difference was higher for the ancestry-specific PRS (LDpred2_EUR_: 0.39 vs LDpred2_AFR_: 0.07, PRSCSX_EUR_: 0.42 vs PRSCSX_AFR_: 0.31) than for the multi-ancestry PRS (LDpred2_combined_EUR_: 0.39 vs Dpred2_combined_AFR_: 0.38, PRSCSX_combined_EUR_: 0.44 vs PRSCSX_combined_AFR_: 0.41). Similarly, among the African-ancestry VTE cases, the mean PRS was higher for the African-ancestry-tuned PRS than for the European-ancestry-tuned PRS, with larger difference for the population-specific PRS (LDpred2_EUR_: 0.18 vs Dpred2_AFR_: 0.19, PRSCSX_EUR_: 0.22 vs PRSCSX_AFR_: 0.28) than the multi-ancestry PRS (LDpred2_combined_EUR_: 0.19 vs Dpred2_combined_AFR_: 0.23, PRSCSX_combined_EUR_: 0.26 vs PRSCSX_combined_AFR_: 0.30).

**Table 1.** Mean and standard deviation of standardized polygenic risk scores with VTE risk in the test set individuals of European and African ancestry.

|  | European |  | African |  |
| --- | --- | --- | --- | --- |
|  | Cases (n=6,781) | Control (n=103,016) | Cases (n=1,385) | Control (n=12,569) |
| Mean (SD) of age at recruitment, in years | 56.9 (13.3) | 52.1 (14.4) | 56.5 (14.6) | 50.8 (16.2) |
| Mean (SD) of LDpred2<sub>EUR</sub> | 0.39 (1.07) | 0 (1) | 0.18 (1.02) | 0 (1) |
| Mean (SD) of LDpred2<sub>AFR</sub> | 0.07 (1) | 0 (1) | 0.19 (1.11) | 0 (1) |
| Mean (SD) of PRSCSX<sub>EUR</sub> | 0.42 (1.07) | 0 (1) | 0.22 (1.03) | 0 (1) |
| Mean (SD) of PRSCSX<sub>AFR</sub> | 0.31 (1.03) | 0 (1) | 0.28 (1.11) | 0 (1) |
| Mean (SD) of LDpred2_combined<sub>EUR</sub> | 0.39 (1.07) | -0.02 (1) | 0.19 (1.02) | 0 (1) |
| Mean (SD) of LDpred2_combined<sub>AFR</sub> | 0.38 (1.06) | 0 (1) | 0.23 (1.04) | 0 (1) |
| Mean (SD) of PRSCSX_combined<sub>EUR</sub> | 0.44 (1.19) | 0 (1) | 0.26 (1.06) | 0 (1) |
| Mean (SD) of PRSCSX_combined<sub>AFR</sub> | 0.41 (1.07) | 0 (1) | 0.3 (1.09) | 0 (1) |

SD, standard deviation; ASN, Asian; EUR, European; PRS, polygenic risk score.

### Evaluation of PRS and VTE risk across populations

**Table 2** shows the estimated OR per SD increase of PRS and AUC for VTE in the test set individuals of European- and African ancestry. For the ancestry-specific PRS, LDpred2_EUR_ and LDpred2_AFR_ were constructed using 604,741 SNPs and 1,184,805 SNPs, respectively, and PRSCSX_EUR_ and PRSCSX_AFR_ were constructed using 591,788 SNPs and 586,660 SNPs, respectively. Multi-ancestry PRS were developed as a linear combination of the ancestry-specific PRS, resulting in 1,212,566 SNPs for LDpred2 and 598,977 SNPs for PRSCSX. The multi-ancestry PRSs outperformed ancestry-specific PRSs in both European- and African-Ancestry test samples and across training methods (LDpred2, PRSCSx) (**Figure 2, S.Figure 1**). In the European-ancestry test set, multi-ancestry PRS in which the weights were tuned in European ancestry samples performed the best (PRSCSX_combined_EUR_: AUC=0.61 (0.6, 0.62), OR=1.48 (1.45, 1.52), LDpred2_combined_EUR_: AUC=0.60 (0.59, 0.61), OR=1.42 (1.39, 1.46)). Similarly, in the African-ancestry test set, a multi-ancestry PRS in which the weights were tuned in African-Ancestry samples performed the best (PRSCSX_combined_AFR_: AUC=0.59 (0.57, 0.60), OR=1.38 (1.30, 1.45); LDpred2_combined_AFR_: AUC=0.57 (0.55, 0.58), OR=1.26 (1.20, 1.33)).

**Figure 2.**
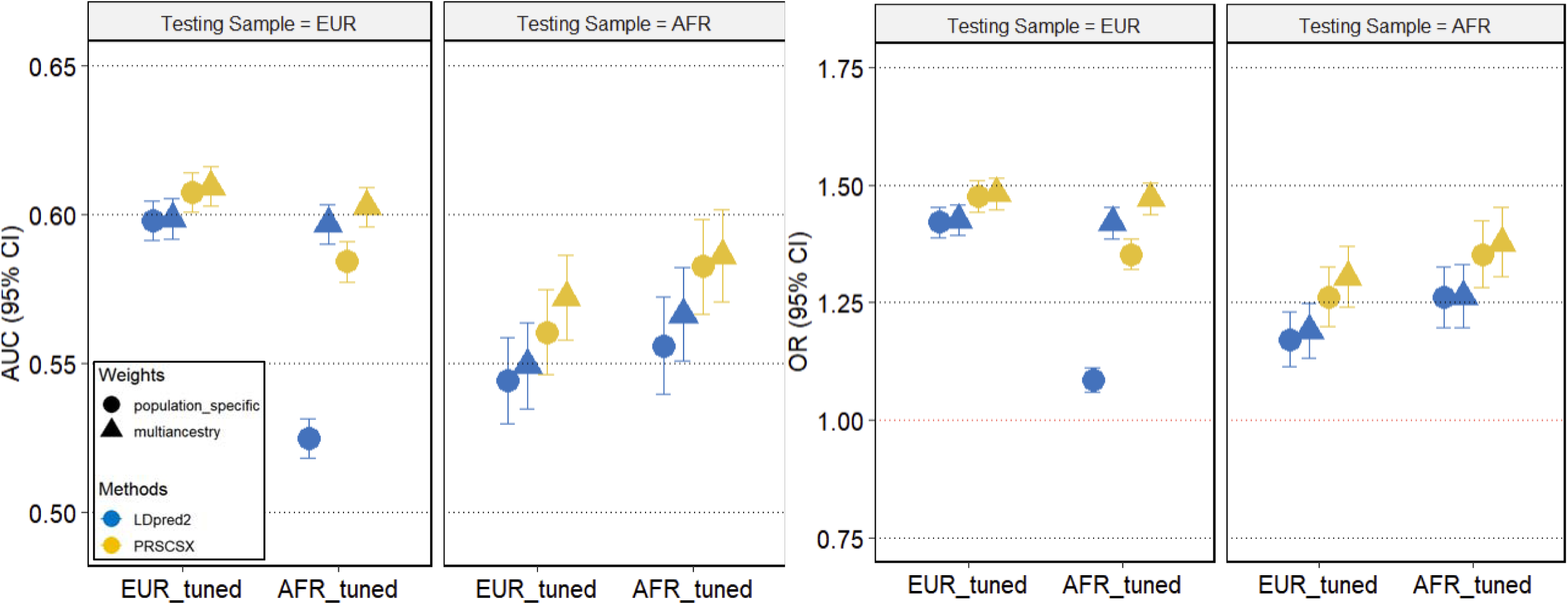
AUC and OR for population-specific and multiancestry PRS across populations.

**Table 2.** Association of polygenic risk scores and VTE risk in the test set individuals of European and African ancestry.

| Method | PRS tuning population | PRS | Number of SNPs | PRS testing population |  |  |  |
| --- | --- | --- | --- | --- | --- | --- | --- |
|  |  |  |  | European |  | African |  |
|  |  |  |  | AUC (95% CI) | Odds ratio per SD (95% CI) | AUC (95% CI) | Odds ratio (95% CI) |
| (1) LDpred2 trained in EUR | - | LDpred2<sub>EUR</sub> | 604,741 | 0.6 (0.59, 0.6) | 1.42 (1.39, 1.45) | 0.54 (0.53, 0.56) | 1.17 (1.11, 1.23) |
| (2) LDpred2 trained in AFR | - | LDpred2<sub>AFR</sub> | 1,184,805 | 0.52 (0.52, 0.53) | 1.09 (1.06, 1.11) | 0.56 (0.54, 0.57) | 1.26 (1.2, 1.33) |
| Combine (1) + (2) | European | <sup>a</sup>LDpred2_combined<sub>EUR</sub> | 1,212,566 | 0.6 (0.59, 0.61) | 1.42 (1.39, 1.46) | 0.55 (0.53, 0.56) | 1.19 (1.13, 1.25) |
| Combine (1) + (2) | African | <sup>a</sup>LDpred2_combined<sub>AFR</sub> | 1,212,566 | 0.6 (0.59, 0.6) | 1.42 (1.39, 1.45) | 0.57 (0.55, 0.58) | 1.26 (1.2, 1.33) |
| (3) PRSCS trained in EUR | - | PRSCSX<sub>EUR</sub> | 591,788 | 0.61 (0.6, 0.61) | 1.47 (1.44, 1.51) | 0.56 (0.55, 0.57) | 1.26 (1.2, 1.33) |
| (4) PRSCS trained in AFR | - | PRSCSX<sub>AFR</sub> | 586,660 | 0.58 (0.58, 0.59) | 1.35 (1.32, 1.39) | 0.58 (0.57, 0.6) | 1.35 (1.28, 1.42) |
| Combine (3) + (4) | European | <sup>a</sup>PRSCSX_combined<sub>EUR</sub> | 598,977 | 0.61 (0.6, 0.62) | 1.48 (1.45, 1.52) | 0.57 (0.56, 0.59) | 1.3 (1.24, 1.37) |
| Combine (3) + (4) | African | <sup>a</sup>PRSCSX_combined<sub>AFR</sub> | 598,977 | 0.6 (0.6, 0.61) | 1.47 (1.44, 1.51) | 0.59 (0.57, 0.6) | 1.38 (1.3, 1.45) |

<sup>a</sup>Combined PRSs were generated using the formula $\alpha_0 + \alpha_1 \text{PRS1} + \alpha_2 \text{PRS2}$ where $\alpha_0$ , $\alpha_1$ and $\alpha_2$ are the weights obtained by fitting a logistic regression model with VTE as outcome, PRS1 and PRS2 as explanatory variables using the validation data set. The weights for the considered combination of PRSs can be found at https://github.com/yonhojee/VTE_PRS.

The association between the PRSs and VTE risk by PRS percentile are shown in **Figure 3**. The association between the highest fifth percentile of PRSCSX_EUR_ (RR=1.89) and LDpred2_EUR_ (RR=1.79) and VTE risk was greater than that of genome-wide significant PRS (RR=1.78). The highest fifth percentile of the best-performing PRS (PRSCSX_combined_EUR_) was associated with 1.9-fold increased risk for VTE among European ancestry subjects compared to the middle stratum (40–50%). Among the African-ancestry samples, the corresponding risk was about 1.68-fold (PRSCSX_combined_AFR_), which is smaller than that in European ancestry samples.

**Figure 3.**
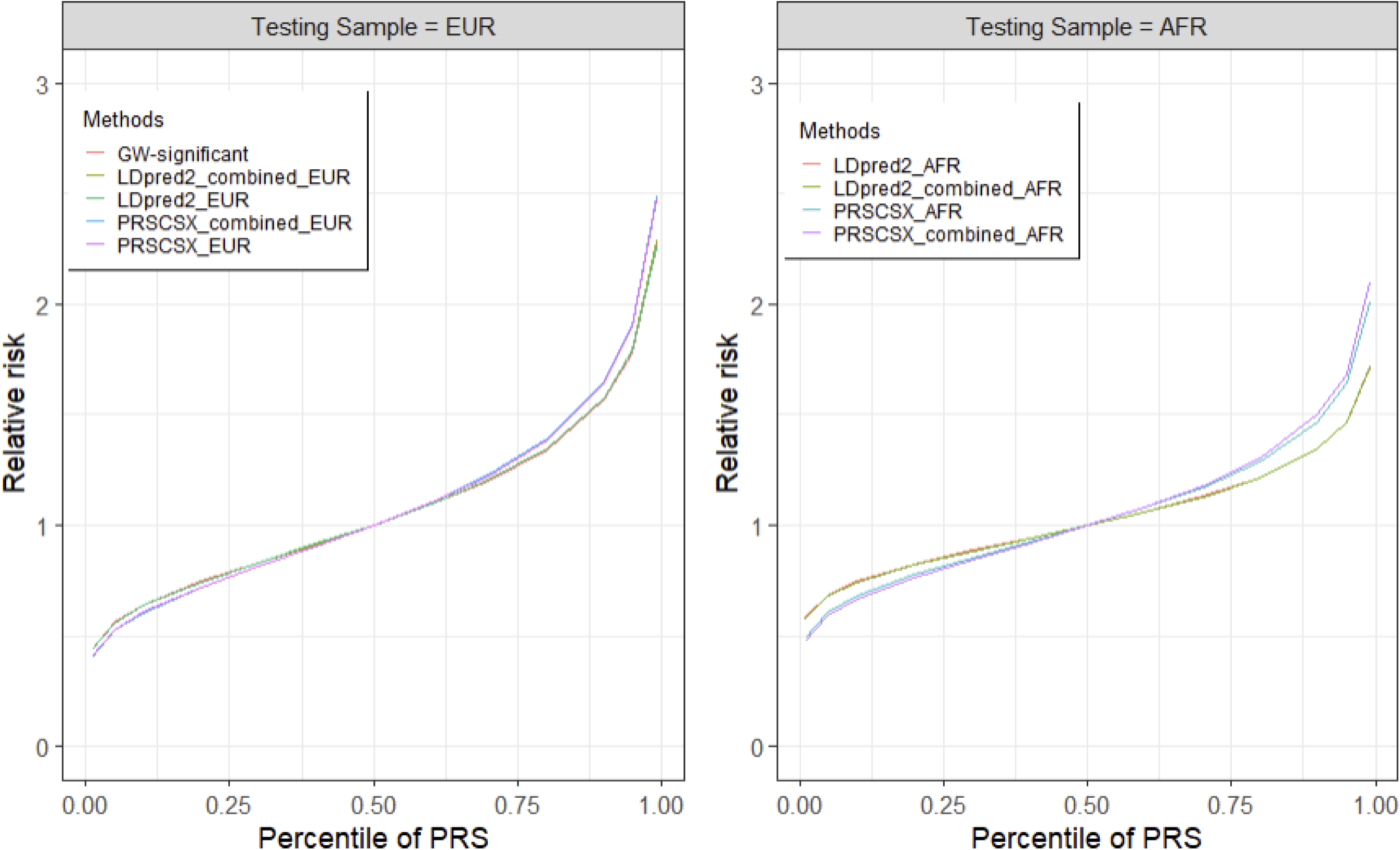
Distribution of relative risk of VTE by PRS across populations.

### Inclusion of known low frequency alleles

When we reconstructed PRS including the five genome-wide significant variants, the new PRS performed worse than our original PRS without the five SNPs in European- (PRSCSX_combined_EUR_: AUC= 0.57 (0.56, 0.59), LDpred2_combined_EUR_: AUC= 0.52 (0.50, 0.53)) and in African-ancestry test samples (PRSCSX_combined_AFR_: AUC= 0.59 (0.58, 0.60), LDpred2_combined_AFR_: AUC= 0.56 (0.55, 0.57)) (**S.Figure 2**). This is likely because the five SNPs are rare in one or both populations (average MAF in European ancestry=0.1, African ancestry=0), and our tuning samples are small, resulting in noisy weights. Future studies with larger and more diverse training samples and further tuning steps are needed to learn better multi-ancestry PRS weights.

## Discussion

Multi-ancestry PRSs outperformed population specific PRSs in U.S. European- and African-ancestry samples, with a greater improvement in African-ancestry samples. The highest fifth percentile of the best performing multi-ancestry PRS in the European ancestry test samples was associated with an approximately 2-fold increased risk for VTE relative to the middle stratum among European-ancestry subjects. The corresponding risk was smaller (1.7-fold) among the African-ancestry subjects, but still non-negligible and higher than any single-ancestry PRS, highlighting that multi-ancestry PRS may be used to identify individuals at highest risk for VTE events. These data may also be useful in guiding primary prevention and treatment strategies across populations, although we stress that demonstrating PRS discrimination is not sufficient to establish clinical utility, which requires consideration of risks and benefits of specific proposed interventions (14,15).

To our knowledge, this is the first attempt to develop PRS of VTE specific to African-ancestry populations. Clinical evaluation of PRS is needed in African-ancestry populations, where the burden of VTE is growing due to its increase in VTE incidence. Our PRS, developed and validated in African-ancestry samples, could be a step towards risk-based clinical management of VTE among Black Americans, as a complement to primary prevention efforts. Black Americans and other population groups suffer social disadvantage and lifestyle risk factors that could be a strong contributors to the disparities in VTE (16). Encouragingly, healthy lifestyle factors were associated with a lower incidence of VTE among people at high genetic risk for VTE (17). Hence, as with most diseases, primary prevention efforts directed at lifestyle interventions to reduce weight or increase activity would have the great potential to reduce the societal burden of VTE. Further research should determine best approaches to VTE prevention that improve health equity.

A recent GWAS meta-analysis demonstrated that European-ancestry individuals at or above the top fifth percentile of a PRS comprised of 37 genome-wide significant variants had a 3.2-fold greater risk for VTE (OR: 3.19; 95% CI: 2.89-3.52) relative to half of the population in the middle of the range (8). More recently, a PRS using the 100 lead variants identified in a larger European ancestry meta-analysis showed AUC=0.620 (95% CI, 0.616–0.625) (9). Since these previous PRS include low MAF variants with large effect sizes (e.g., rs6025: transancestry OR=2.39 (8) on *F5* gene), the performance of these previous PRSs and our PRSs is not directly comparable. It is worth noting that our PRS was built using genome-wide common variants and was designed to be transportable between European- and African-ancestry individuals, which can be useful for settings with diverse genetic background. The PRSs presented here complement the low-frequency, large-effect variants and clinical and behavioral risk factors; future work should develop and evaluate comprehensive risk models combining multi-ancestry PRS, low-frequency variants and other risk factors.

The major strength of the study is that it is the first attempt to develop and validate multi-ancestry PRS for VTE, providing potential utility of PRS in VTE prevention among African-ancestry populations, where the VTE burden is high. In addition, we validated the PRS in the five independent biobanks from GBMI using harmonized analysis framework (e.g. phenotype definitions, ancestry assignments, and PRS construction).

There are several limitations in our study. First, we have focused on common SNPs, specifically HapMap3 SNPs for VTE PRS construction. As a result, information from rarer variants missing in the LD reference panel may not be captured in other non-European ancestries. Second, the lower predictive ability of VTE PRS in African-ancestry samples can be explained by smaller sample size of African-ancestry VTE meta-analysis GWAS, which is 10 times smaller than European GWAS. Third, there remains a multitude of factors that may contribute to cross-biobank heterogeneity including phenotype precision, cohort-level disease prevalence, and environmental factors. We have provided analysis results by cohort (**Supplementary Figure 1**).

## Conclusions

We found that multi-ancestry PRS for VTE outperformed population-specific PRS, especially in African ancestry populations with relatively small GWAS sample sizes. These findings suggest that the multi-ancestry PRS may be used to identify individuals at highest risk for VTE event and provide guidance for the most effective treatment strategy across populations.

## Materials and Methods

### Study populations

We trained the PRS using summary statistics from the International Network against Venous Thrombosis (INVENT) consortium cross-ancestry GWAS meta-analyses of European- (71,771 VTE cases and 1,059,740 controls) and African-ancestry samples (7,482 VTE cases and 129,975 controls) (9). The meta-analysis is based on prospective cohorts and case-control data from 30 studies.

Tuning (1,329 cases and 1,324 controls of European-ancestry and 238 cases and 3,589 controls of African-ancestry) and validation data (6,781 cases and 103,016 controls of European ancestry and 1,385 cases and 12,569 controls of African ancestry) came from Nurses’ Health Study [NHS] and Health Professional Follow-up Study [HPFS] and 4 Global Biobank Meta-analysis Initiative (GBMI) biobanks (Michigan Genomics Initiative [MGI], UCLA Precision Health Biobank [UCLA], Penn Medicine Biobank [PMBB], and Lifelines) with representation across African and European-ancestry populations included (**Figure 1**). These tuning and validation data were not included in the GWAS used in the training step. The definitions of African- and European-ancestry populations in each study are provided in the **Supplementary Materials**; these definitions typically involve both self-reported race and ethnicity and genetic similarity to a set of (study-specific) labeled reference samples.

**Supplementary Table 1** summarizes the study design, genotyping arrays, and the sample size in each study. All studies were approved by the relevant institutional ethics committees and review boards, and all participants provided written informed consent.

### Statistical methods

#### PRS training and tuning

##### PRS training and tuning using LDpred2

We ran LDpred2-auto(14) to construct PRS on HapMap3 variants using the INVENT GWAS meta-analysis summary statistics corresponding to each population. We constructed linkage disequilibrium (LD) reference panels for the development of the European-ancestry PRS (LDpred2_EUR_) and African-Ancestry PRS (LDpred2_AFR_) using the EUR and AFR supersamples from the 1000 Genomes Project (Phase 3), respectively.(18) These population-specific PRSs were then linearly combined to construct multi-ancestry PRS (LDpred2_EUR_ + LDpred2_AFR_) in which the relative contribution of each PRS was estimated by logistic regression in the tuning dataset of European-ancestry samples (LDpred2_combined_EUR_) and African-ancestry samples (LDpred2_combined_AFR_). Analyses were run using R; code is available at https://github.com/yonhojee/VTE_PRS.

##### PRS training and tuning using PRSCSx

We separately applied PRSCSx(15) to the summary statistics from the European- and African-ancestry INVENT VTE GWAS, using the EUR and AFR LD reference panels from the 1000 Genomes Project (Phase 3). The global shrinkage parameter was learnt from the data using a fully Bayesian approach. Ancestry-specific PRSs generated using European- (PRSCSx_EUR_) and African-specific posterior weights (hereafter denoted as PRSCSx_AFR_) were linearly combined to construct multi-ancestry PRS (PRSCSx_EUR_ + PRSCSx_AFR_). The regression coefficients for the linear combination were obtained by fitting a logistic regression model in the tuning data set of European ancestry samples (PRSCSx_combined_EUR_) and African American samples (PRSCSx_combined_AFR_). Analyses were run using Python; code is available at https://github.com/yonhojee/VTE_PRS.

#### PRS validation

In each test dataset, population-specific PRSs were calculated as 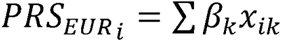 and 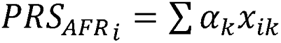, where *x_ik_* is the dosage of risk allele (0–2) at genetic variant *k* for subject *i*, and *β_k_* and *α_k_* are the corresponding weight in European and African PRS, respectively. The estimates of *β_k_* and *α_k_* were trained using summary statistics from the INVENT consortium and LDpred2 and PRSCSx as described above.

We calculated the multi-ancestry PRSs as the linear combination of European- and African-ancestry specific PRS:

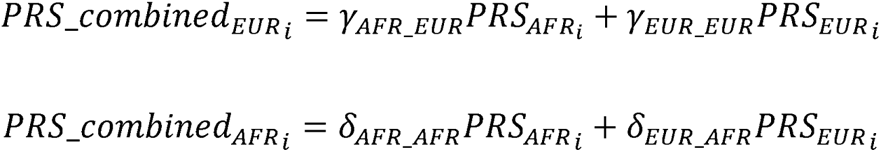

where PRS_AFR_ and PRS_EUR_ are the PRSs trained in single-ancestry GWAS and the *γ* and *δ* are “meta-weights” tuned in European- and African-ancestry samples, respectively. SNPs with imputation R^2^ > 0.9 in training dataset were retained for subsequent analyses. The lists of SNPs and the weights for the PRS computation are available at https://github.com/yonhojee/VTE_PRS.

PRSs were standardized within each validation sample to have unit SD in the control subjects. Logistic regression, adjusting for ten principal components and sex, was used to estimate odds ratios (ORs) for association between the standardized PRSs and VTE risk in each testing set. The discrimination of PRS was assessed using area under the receiver operating curve (AUC). The OR per SD and AUC were obtained individually for each study and combined separately for European- and African-ancestry samples using a fixed-effect meta-analysis.

All statistical analyses were conducted using R v.4.3.0. Logistic regression and AUC were done using *glm()* and *roc()* in R.

#### The distribution of relative risk of VTE by PRS across populations

We simulated 100,000 individuals with PRS distribution of N(0,1) multiplied by log OR per SD estimates for each PRS. The simulated PRS was then exponentiated to estimate relative risk estimates and split into the percentile categories: [0–1%] (1-5%], (5-10%], (10–20%], (20–30%], (30–40%], (40–50%] (reference group), (50–60%], (60–70%], (70–80%], (80–90%], (90–95%], (95–99%] and (99–100%].

#### Sensitivity analysis of including known low frequency alleles

Out of the 37 genome-wide significant variants, our current PRSs do not include five variants (rs6025, rs145470028, rs1799963, rs6048, and rs143478537), which would have been filtered out of our analyses for one reason or another (e.g., on the X chromosome, low minor allele frequency [MAF]). These variants are important to be considered in VTE PRS given their large effect sizes (e.g., rs6025: transancestry OR=2.39(8) on *F5* gene). As a sensitivity analysis, we constructed new PRSs, which additionally include these previously reported variants that are i) not included in our PRS due to the low frequency and ii) not in LD with the variants already included in our PRS. The final PRSs were obtained by the linear combination of the original PRS (constructed using common variants only) and the additional SNPs where the coefficients for the original PRS and the additional SNPs were tuned in the independent ancestry-specific samples (See more details in the **Supplementary Materials**).

## Data Availability

All data produced are available online at https://github.com/yonhojee/VTE_PRS.

## Acknowledgements

The views expressed in this manuscript are those of the authors and do not necessarily represent the views of the National Cancer Institute; National Heart, Lung, and Blood Institute; the National Institutes of Health; or the US Department of Health and Human Services.

This work was supported by the National Institutes of Health grant 5U01CA261339.

The INVENT Consortium acknowledges all of the participants across the studies who provided their health information to support these analyses.

The authors would like to thank the participants and staff of the NHS and NHSII for their valuable contributions. The NHS, NHS-II and HPFS were supported by grants UM1 CA186107, U01 CA176726, and U01 CA167552 from the National Institutes of Health.

The authors acknowledge the Michigan Genomics Initiative participants, Precision Health at the University of Michigan, the University of Michigan Medical School Central Biorepository, and the University of Michigan Advanced Genomics Core for providing data and specimen storage, management, processing, and distribution services, and the Center for Statistical Genetics in the Department of Biostatistics at the School of Public Health for genotype data curation, imputation, and management in support of the research reported in this publication.

We gratefully acknowledge the resources provided by the Institute for Precision Health (IPH) and participating UCLA ATLAS Community Health Initiative patients. The UCLA ATLAS Community Health Initiative in collaboration with UCLA ATLAS Precision Health Biobank, is a program of IPH, which directs and supports the biobanking and genotyping of biospecimen samples from participating UCLA patients in collaboration with the David Geffen School of Medicine, UCLA CTSI and UCLA Health. Members of the UCLA ATLAS Community Health Initiative include Ruth Johnson, Yi Ding, Vidhya Venkateswaran, Arjun Bhattacharya, Alec Chiu, Tommer Schwarz, Malika Freund, Lingyu Zhan, Kathryn S. Burch, Christa Caggiano, Brian Hill, Nadav Rakocz, Brunilda Balliu, Jae Hoon Sul, Noah Zaitlen, Valerie A. Arboleda, Eran Halperin, Sriram Sankararaman, Manish J. Butte, Clara Lajonchere, Daniel H. Geschwind, and Bogdan Pasaniuc, on behalf of the UCLA Precision Health Data Discovery Repository Working Group and UCLA Precision Health ATLAS Working Group.

We acknowledge the PMBB for providing data and thank the patient-participants of Penn Medicine who consented to participate in this research program. We would also like to thank the Penn Medicine BioBank team and Regeneron Genetics Center for providing genetic variant data for analysis. The PMBB is approved under IRB protocol# 813913 and supported by Perelman School of Medicine at University of Pennsylvania, a gift from the Smilow family, and the National Center for Advancing Translational Sciences of the National Institutes of Health under CTSA award number UL1TR001878.

The Lifelines Biobank initiative has been made possible by funding from the Dutch Ministry of Health, Welfare and Sport, the Dutch Ministry of Economic Affairs, the University Medical Center Groningen (UMCG the Netherlands), University of Groningen and the Northern Provinces of the Netherlands. The generation and management of GWAS genotype data for the Lifelines Cohort Study is supported by the UMCG Genetics Lifelines Initiative (UGLI). UGLI is partly supported by a Spinoza Grant from NWO, awarded to Cisca Wijmenga. The authors wish to acknowledge the services of the Lifelines Cohort Study, the contributing research centers delivering data to Lifelines, and all the study participants.

UGLI Lifelines Cohort Study group author: Raul Aguirre-Gamboa (1), Patrick Deelen (1), Lude Franke (1), Jan A Kuivenhoven (2), Esteban A Lopera Maya (1), Ilja M Nolte (3), Serena Sanna (1), Harold Snieder (3), Morris A Swertz (1), Peter M. Visscher (3,4), Judith M Vonk (3), Cisca Wijmenga (1), Naomi Wray (4); (1) Department of Genetics, University of Groningen, University Medical Center Groningen, The Netherlands; (2) Department of Pediatrics, University of Groningen, University Medical Center Groningen, The Netherlands; (3) Department of Epidemiology, University of Groningen, University Medical Center Groningen, The Netherlands; (4) Institute for Molecular Bioscience, The University of Queensland, Brisbane, Queensland, Australia.

