## Supplementary Materials for "Multi-ancestry polygenic risk scores for venous thromboembolism"

**SUPPLEMENTARY METHODS**

**Participating Studies**

The current cross-ancestry PRS validation study is comprised of new analyses of data from 5 studies, including the Nurses’ Health Study I and II (NHS and NHSII) and Health Professional Follow-up Study (HPFS), Michigan Genomics Initiative (MGI), UCLA Precision Health Biobank (UCLA), Penn Medicine Biobank (PMBB), and Lifelines as well as previously published data from the INVENT consortium, a 17 study analysis of prospective cohorts and case-control data (designated INVENT-2019) ^1^.

**Genotyping and Imputation**

Genotyping was performed with high-density genome-wide arrays for all studies. Genotype data were imputed to the 1000 Genomes Project reference dataset.

**Phenotype definitions (recommended by** [**GBMI**](https://www.medrxiv.org/content/10.1101/2021.11.19.21266436v1)**):**

- Controls (subjects with these ICD codes are excluded from controls): I81, I82.0, I80.0, I80.3, I80.8, I80.9, D68, 452,453.0,451.0,451.89,451.9,286
- Cases: I80.1, I80.2, I82.2, I26.0, I26.9, 451.11, 453.40, 453.2, 453.77, 453.87, 415.1
- OPCS codes to define cases: L79.1, L90.2

**PRS with additional five genome-wide significant variants**

Previously reported genome-wide significant SNPs^2^ that are not included in the PRS and not in LD with the SNPs in the PRS were additionally included using weights from INVENT GWAS. The final PRSs were obtained by the linear combination of the original PRS (-temp) and additional SNPs (_ADD) where the coefficients for each term (b1, b2) were tuned in the independent ancestry-specific samples.

Step 1. Compute “population-specific PRS-temp” under the pre-specified quality control (SNPs with imputation R^2^ > 0.9)

- $PR{S_{EUR}}_{i}= \sum\beta_{k}x_{ik}$ and $PR{S_{AFR}}_{j}= \sum\alpha_{k}y_{jk}$*,*

where *x_ik_* and *y_jk_* are the dosage of risk allele (0-2) at SNP *k* for an European subject *i* and African subject *j,* respectively, $\beta_{k}$ and $\alpha_{k}$ are the corresponding weight in European and African PRS, respectively.

Step 2. Compute “PRS-ADD” consisting of SNPs that are either not included or not in LD with the SNPs in the EUR and AFR PRS-temp.

For biobanks with chromosome X data

- EUR_ADD = 0.6893*rs1799963+0.0637*rs6048+-0.0697*rs143478537
- AFR_ADD = 1.0864*rs6025+-0.058*rs145470028 + 0.6758*rs1799963 + 0.0943*rs6048+-8e-04*rs143478537

For biobanks without chromosome X data

- EUR_ADD = 0.6893*rs1799963
- AFR_ADD = 1.0864*rs6025+-0.058*rs145470028 + 0.6758*rs1799963

Step 3. Generate the final population-specific PRS: linear combination of the PRS-temp and PRS-ADD, where the coefficients for each term (b1, b2) were tuned in the independent ancestry-specific samples.

For biobanks with chromosome X data

- LDPRED2_EUR = LDPRED2_EUR_temp*0.7843475 + EUR_ADD*0.2560768
- LDPRED2_AFR = LDPRED2_AFR_temp*0.6531995 + AFR_ADD*1.0464763
- PRSCSX_EUR = PRSCSX_EUR_temp*5.2481014 + EUR_ADD*0.3647353
- PRSCSX_AFR = PRSCSX_AFR_temp*1.1766993 + AFR_ADD*0.9679427

For biobanks without chromosome X data

- LDPRED2_EUR_NEW = LDPRED2_EUR_temp*0.7846153 + EUR_ADD*0.3881513
- LDPRED2_AFR_NEW = LDPRED2_AFR_temp*0.6243749 + AFR_ADD*1.0810380)
- PRSCSX_EUR_NEW = PRSCSX_EUR_temp*5.2476172 + EUR_ADD*0.5289784
- PRSCSX_AFR_NEW = PRSCSX_AFR_temp*1.1475613 + AFR_ADD*0.9967247

Step 4. Generate the final multi-ancestry PRS: linear combination of population-specific PRS-temp and a weighted sum of SNPs that do not appear in any of the temp PRS in the regression

For biobanks with chromosome X data

- LDpred2_combined_EUR = LDPRED2_EUR_temp*0.7846796 + LDPRED2_AFR_temp*1.1920742 + EUR_ADD*0.2425207
- LDpred2_combined_AFR = LDPRED2_EUR_temp*0.3815917 + LDPRED2_AFR_temp*0.7126167 + EUR_ADD*(-0.9902129)
- PRSCSX_combined_EUR = PRSCSX_EUR_temp*4.7125752 + PRSCSX_AFR_temp*0.6508282 + EUR_ADD*0.3762028
- PRSCSX_combined_AFR = PRSCSX_EUR_temp*2.7844363 + PRSCSX_AFR_temp*0.9204827 + EUR_ADD*(-0.8456485)

For biobanks without chromosome X data

- LDpred2_combined_EUR_NEW (European weight tuned) = LDPRED2_EUR_temp*0.7849432 + LDPRED2_AFR_temp*1.2121711 + EUR_ADD*0.3816551
- LDpred2_combined_AFR_NEW (African weight tuned) = LDPRED2_EUR_temp*0.3834051 + LDPRED2_AFR_temp*0.7497259 + EUR_ADD*(-19.0491483)
- PRSCSX_combined_EUR_NEW (European weight tuned) = PRSCSX_EUR_temp*4.7118682 + PRSCSX_AFR_temp*0.6511214 + EUR_ADD*0.5561337
- PRSCSX_combined_AFR_NEW (African weight tuned) = PRSCSX_EUR_temp*2.7621930 + PRSCSX_AFR_temp*0.9572978 + EUR_ADD*(-18.8000576)

**STUDY DESCRIPTIONS**

The **Nurse’s Health Studies (NHS and NHS-II)** These studies have been described previously and additional information is available at <http://www.nurseshealthstudy.org> ^3,4^. NHS and NHS-II are longitudinal cohort studies of female nurses. In 1976, baseline questionnaires were sent to registered nurses from 11 populous US states, establishing a cohort of 121,700 women aged 30-55. There were no exclusions by race, but the majority (96%) were of European ancestry; corresponding to the demographics of nurses in 1976. NHS participants are mailed a questionnaire every two years that assesses risk factor status and interval disease events. Physician-diagnosed PE has been asked on every biennial NHS questionnaire since 1982. In NHS, the question reads: “Since [year], have you had any of these physician-diagnosed illnesses? ... Pulmonary Embolus.” NHS questionnaires also ask whether the nurse had “Any other major diagnosis: ____.” In NHS, DVT is captured when a nurse answers that she has had phlebitis or thrombophlebitis (ICD-9=453.x). Questionnaire-reported VTE diagnoses have proven to be highly accurate, with >95% validation of VTE events. A physician reviews medical records for all reported PEs, validating diagnoses when medical records include: a positive pulmonary angiogram, a high-probability ventilation/perfusion scan, or a positive CT pulmonary angiogram.

The **Health Professional Follow-up Study (HPFS)** has been described previously and additional information is available at <https://sites.sph.harvard.edu/hpfs/> ^5^. HPFS was started in 1986 as a companion study to the Nurses’ Health Study, enrolling 51,529 male dentists, optometrists, osteopathic physicians, pharmacists, podiatrists and veterinarians 40-75 years old. There were no exclusions by race, but the majority of participants were of European ancestry; corresponding to the demographics of health professionals in 1986. HPFS participants are mailed a questionnaire every two years that assesses risk factor status and interval disease events. In HPFS, DVT and PE are captured together by the question: “Since [year], have you had any of these physician-diagnosed illnesses? Deep Vein Thrombosis/Pulmonary Embolus”. Questionnaire-reported VTE diagnoses have proven to be highly accurate, with >95% validation of VTE events. A physician reviews medical records for all reported PEs, validating diagnoses when medical records include: a positive pulmonary angiogram, a high-probability ventilation/perfusion scan, or a positive CT pulmonary angiogram.

The ancestry information fir NHS, NHS-II, and HPFS was obtained by self-reported as European or African ancestry and clustered with other European or African ancestry samples according to principal components.

The **Michigan Genome Initiative (MGI)** has been described previously^6^ and additional information is available at <https://precisionhealth.umich.edu/our-research/michigangenomics/>. MGI is a single-health-system biobank consisting of more than 91,000 participants recruited primarily during surgical encounters at Michigan Medicine, the University of Michigan’s healthcare system ^6^. Recruitment for MGI started in 2012 and has primarily occurred through the Department of Anesthesiology during inpatient surgical procedures at Michigan Medicine. Recruiting during a surgical encounter provides a convenient opportunity to obtain patient consent, complete questionnaires, and collect a blood sample biospecimen. In this analysis, we use the genetic and clinical data for MGI “Freeze 3” (March 23, 2020) comprised of 57,055 participants. Individuals from Freeze 3 were excluded if they had the ICD codes: I81, I82.0, I80.0, I80.3, I80.8, I80.9, D68, 452, 453.0, 451.0, 451.89, 451.9, 286. This left 55,049 total individuals in the study comprising of 51,222 Europeans and 3,827 African Americans.

The **UCLA Precision Health Biobank (UCLA)** has been described previously ^7^ and is a biobanking initiative within a larger academic medical center. The UCLA Health system includes 2 hospitals and a total of 210 primary and specialty outpatient locations located primarily in the greater Los Angeles area. The UCLA Health system serves approximately 5% of Los Angeles County population. An electronic form of health records was implemented throughout the UCLA Health system in 2013 and a variety of longitudinal clinical information has been identified and made available to researchers. The average age of participants, defined as a participant’s age recorded in the EHR as of September 2021, is 55.6 (SD: 17.2) years with an average medical record length of 11.6 (SD: 8.5) years. Individuals opting in to the biobank have more healthcare interactions than the general population as has been previously described.

For genetic ancestry determination we used PCA as described in Johnson et al 2023^7^: PCA was performed on a merged dataset consisting of individuals from ATLAS merged with individuals from the 1000 Genomes Project reference panel. This reference panel consists of genotypes from individuals of known European, African, admixed American, East Asian, and South Asian descent. After projecting the PCs into two-dimensional space, the labeled samples were used from 1000 Genomes to define cluster boundaries for individuals in ATLAS corresponding to each continental ancestry group. Cluster thresholds were visually determined by comparing the overlap of the 1000 Genomes reference panel samples with ATLAS samples in PC space.

The **Penn Medicine BioBank (PMBB)** is a research program that recruits patients from the University of Pennsylvania Health System for genomic and precision medicine research. The PMBB researchers have access to EHR data linked with genetic data from the biobank ^8^. Currently, the PMBB has over 2,000,000 participants enrolled and whole exome sequencing and genotyping is available on 43,623 patient participants. However, a subset of approximately 6,591 subjects were excluded from this analysis as they were used in previous genome-wide association studies (GWAS) of venous thromboembolism (VTE) from INVENT consortium ^1^. The PMBB is one of the most diverse medical biobanks, with approximately 30% of participants of non-European ancestry. The EHR data in the PMBB includes a median of seven years of longitudinal data. This analysis was conducted on 33,1000 samples including 3,399 VTE cases and 29,701 controls. The cases included 1,708 (50.2%) males and 1,691 (49.2%) females while the controls included 14,125 (47.6%) males and 15,566 (52.4%). Ancestry was defined by genetic ancestry whereby patients with PCs mapping to African ancestry and European ancestry defined by the 1000 Genomes Project were denoted as being of African and European ancestry respectively. For the purposes of this study, cases contained 2,000 patients with European ancestry and 1,399 patients with African ancestry, while controls contained 20,460 patients with European ancestry and 10,792 patients with African ancestry.

The **Lifelines** is a multi-disciplinary prospective population-based cohort study examining in a unique three-generation design the health and health-related behaviours of 167,729 persons living in the North of Netherlands. It employs a broad range of investigative procedures in assessing the biomedical, socio-demographic, behavioural, physical and psychological factors which contribute to the health and disease of the general population, with a special focus on multi-morbidity and complex genetics. Participants completed questionnaires, underwent physical examinations, and biological samples including DNA were collected^9^. The majority (>99%) of Lifelines participants are of European ancestry. Venous thromboembolism (VTE) phenotype was defined using a classification based on questions about self-reported thrombosis events, thrombosis-specific medications, thrombosis-specific complaints and for this study, we included those for whom genotyping data were available. Controls in this context were those individuals who were classified as being free of thrombosis and related symptoms, and for whom genotyping data were available.

The Lifelines Biobank initiative has been made possible by funding from the Dutch Ministry of Health, Welfare and Sport, the Dutch Ministry of Economic Affairs, the University Medical Center Groningen (UMCG the Netherlands), University of Groningen and the Northern Provinces of the Netherlands. The generation and management of GWAS genotype data for the Lifelines Cohort Study is supported by the UMCG Genetics Lifelines Initiative (UGLI). UGLI is partly supported by a Spinoza Grant from NWO, awarded to Cisca Wijmenga. The authors wish to acknowledge the services of the Lifelines Cohort Study, the contributing research centers delivering data to Lifelines, and all the study participants.

Using genotype data from Lifelines, 1000 Genomes (1000G) and GoNL individuals, a combined dataset was created with markers present in all three datasets. SNPs in this combined dataset were filtered by MAF > 10%, call rate > 99% and HLA region removed. This dataset was further pruned. Principal components were then computed using only the participants from the 1000G and GoNL datasets and projecting the Lifelines participants into them. Next, the first two PCs were used to flag as European all individuals clustering with GoNL or 1000G European populations or no more than 3 SD away from their extremes according to both PC1 and PC2.

UMCG Genetics Lifelines Initiative (UGLI) group author: LifeLines Cohort Study

Raul Aguirre-Gamboa (1), Patrick Deelen (1), Lude Franke (1), Jan A Kuivenhoven (2), Esteban A Lopera Maya (1), Ilja M Nolte (3), Serena Sanna (1), Harold Snieder (3), Morris A Swertz (1), Peter M. Visscher (3,4), Judith M Vonk (3), Cisca Wijmenga (1), Naomi Wray (4)

(1) *Department of Genetics, University of Groningen, University Medical Center Groningen, The Netherlands*
(2) *Department of Pediatrics, University of Groningen, University Medical Center Groningen, The Netherlands*
(3) *Department of Epidemiology, University of Groningen, University Medical Center Groningen, The Netherlands*
(4) *Institute for Molecular Bioscience, The University of Queensland, Brisbane, Queensland, Australia.*

**Supplementary Figure 1**. Association between PRS and VTE risk across participating studies.

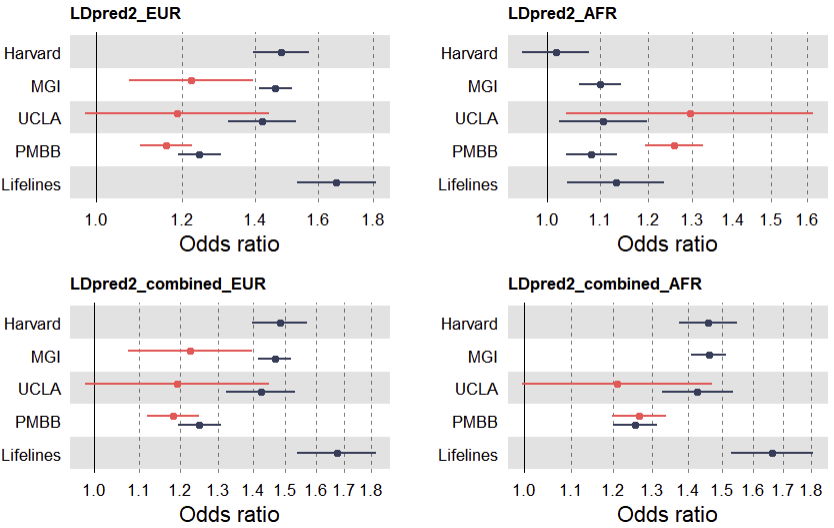

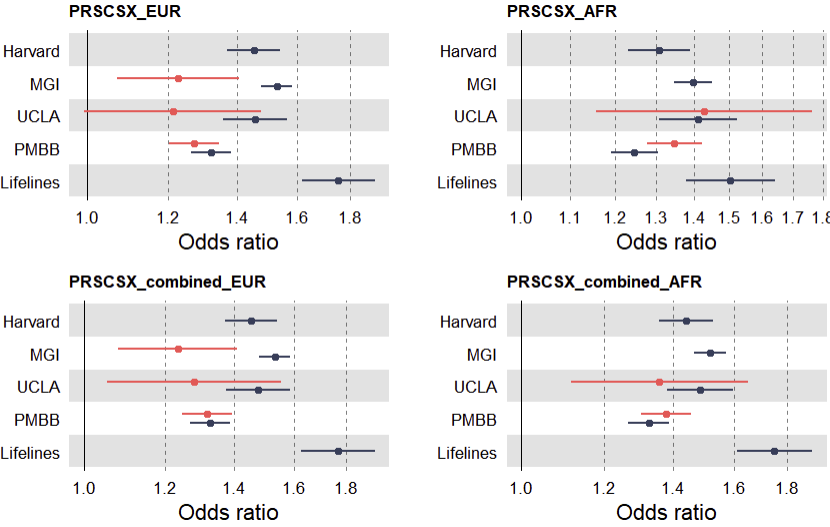

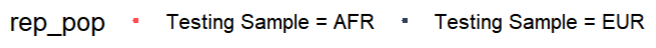

**Supplementary Figure 2**. AUC and OR for population-specific and multiancestry PRS with additional genome-wide significant variants across populations.

Biobanks with chromosome X

- EUR: Harvard, MGI, Lifelines, PMBB
- AFR: MGI (EUR_tuned), PMBB

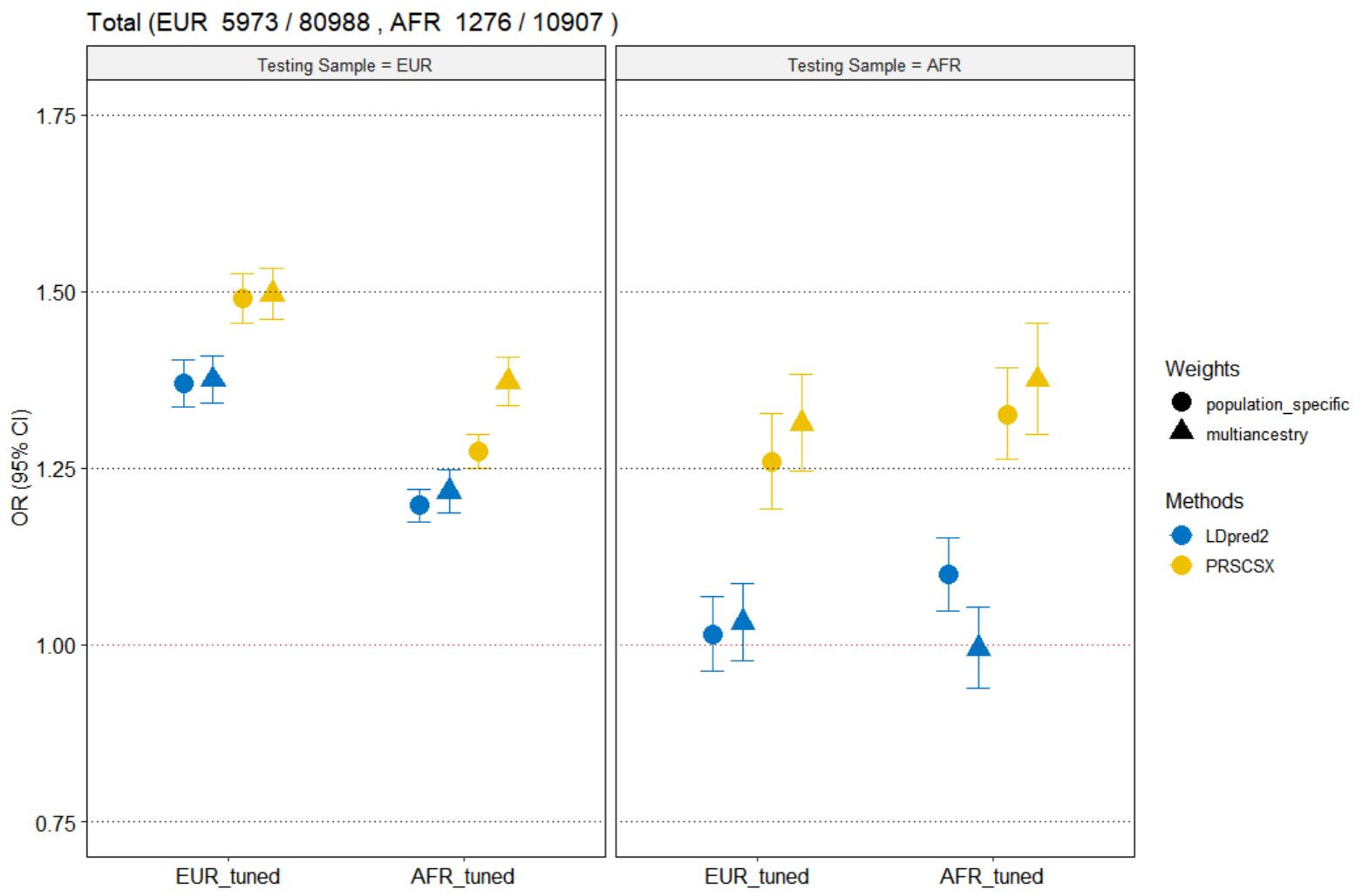

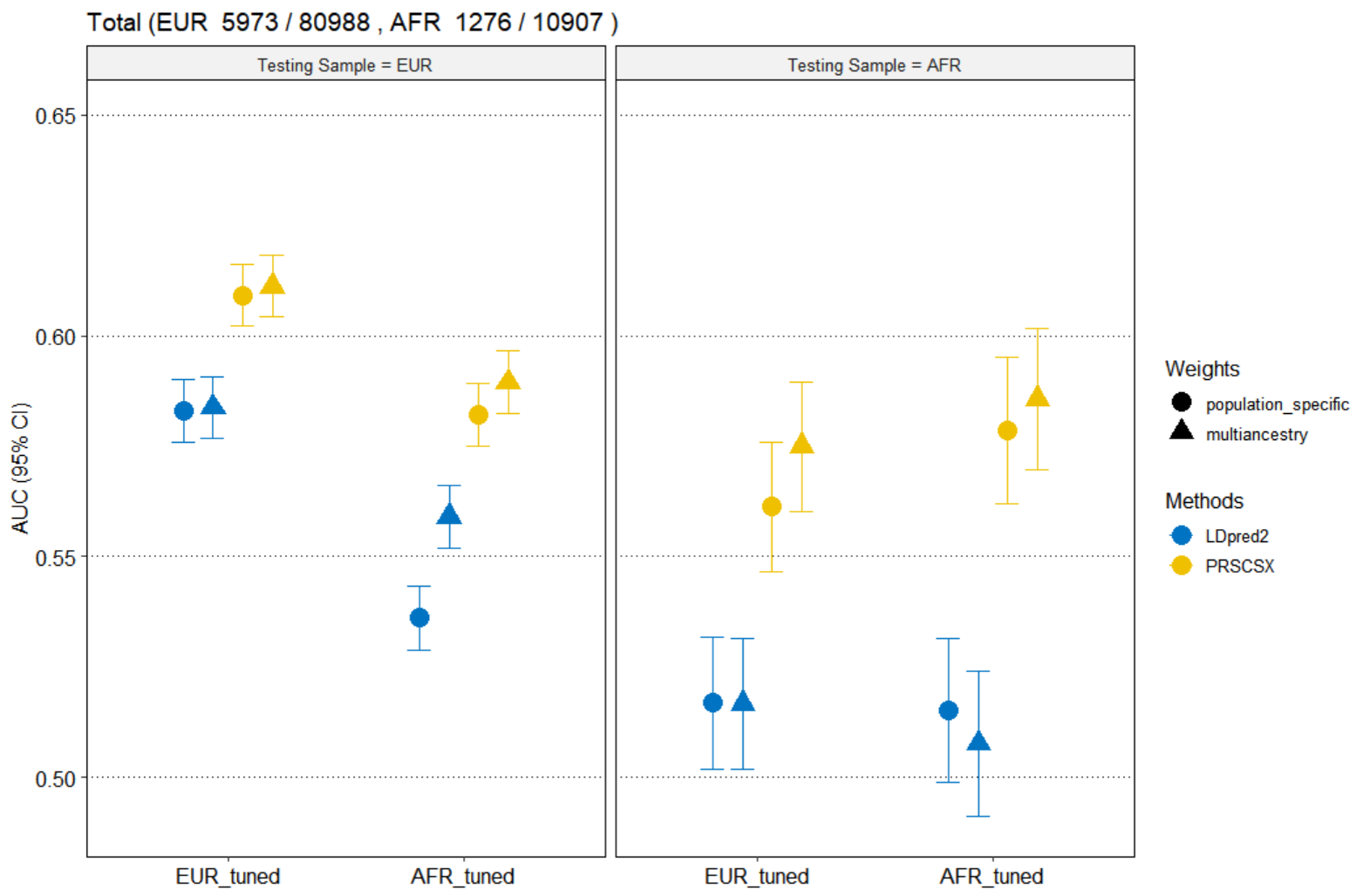

Biobanks without chromosome X

- EUR: Harvard, MGI, Lifelines, PMBB, UCLA
-
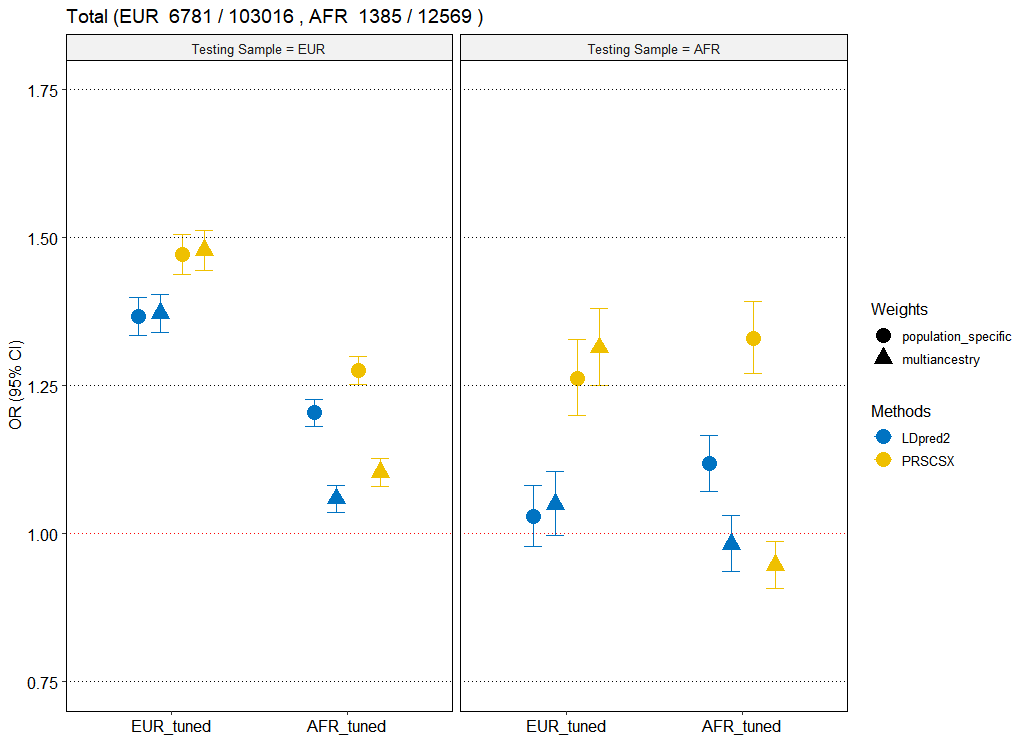

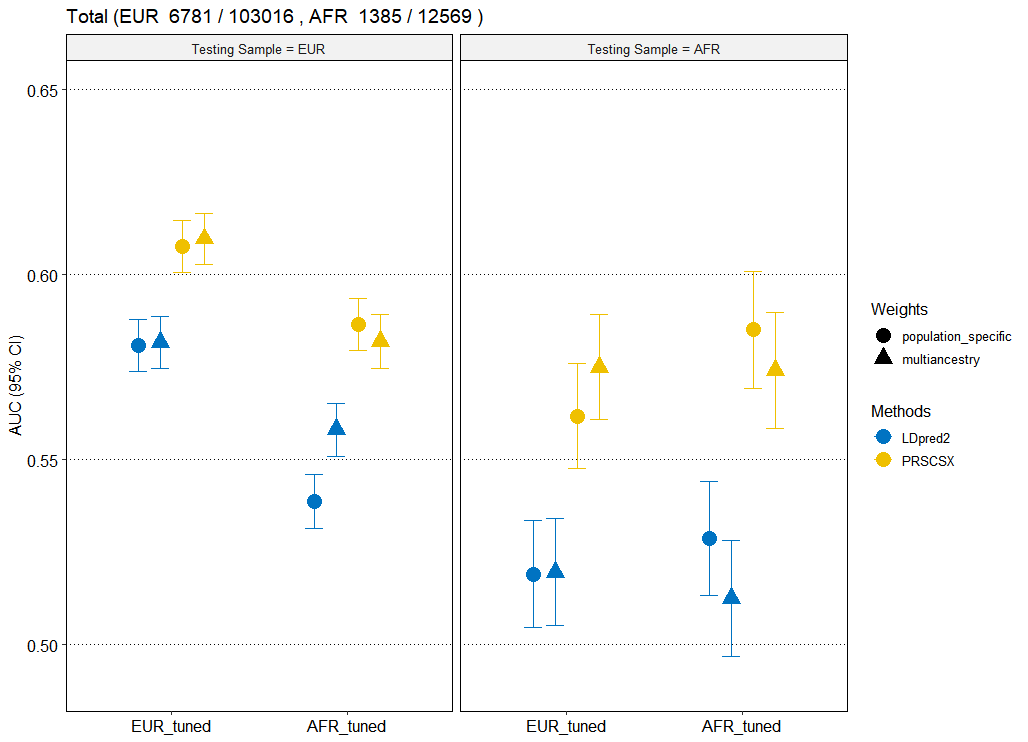

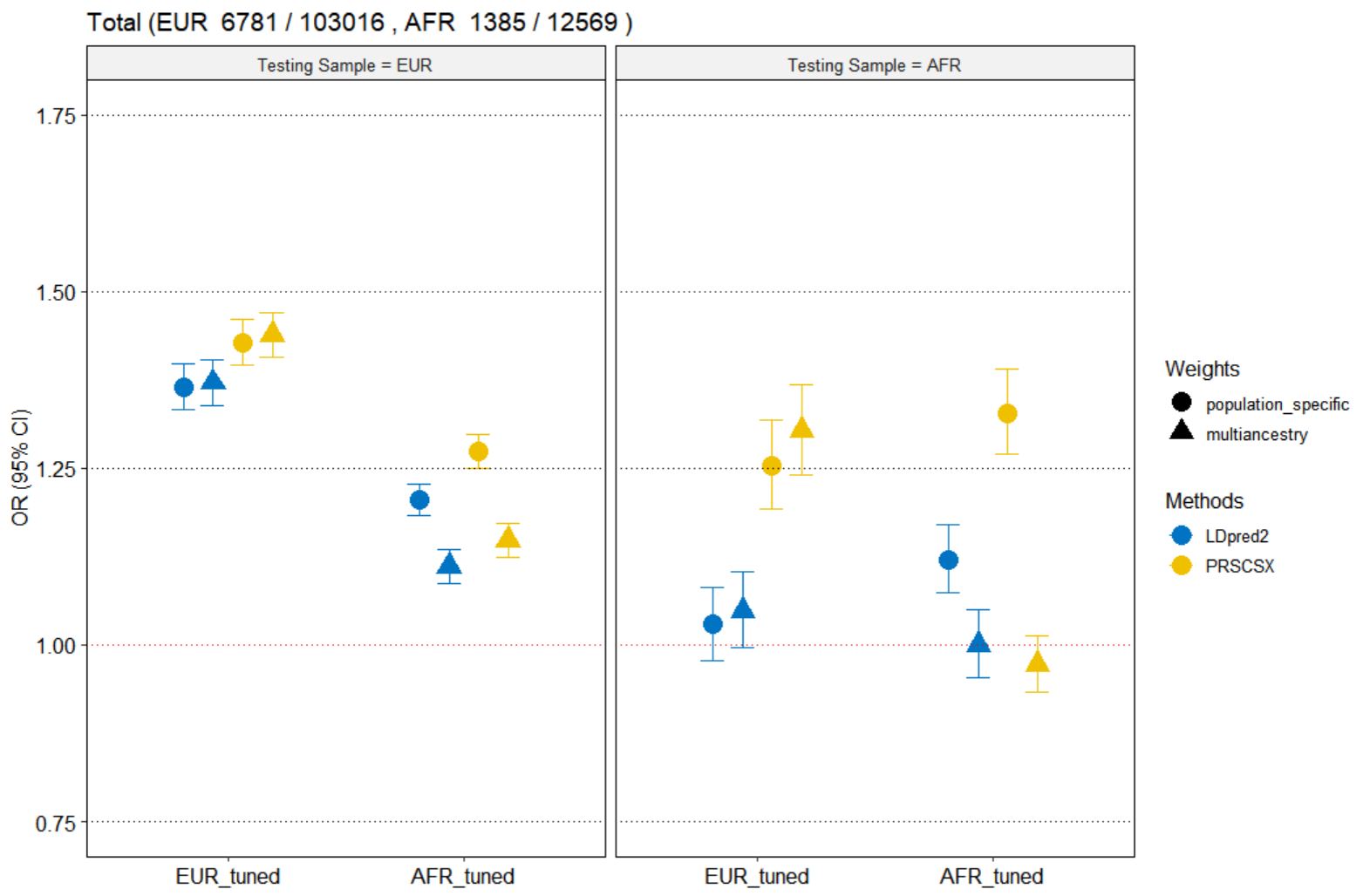

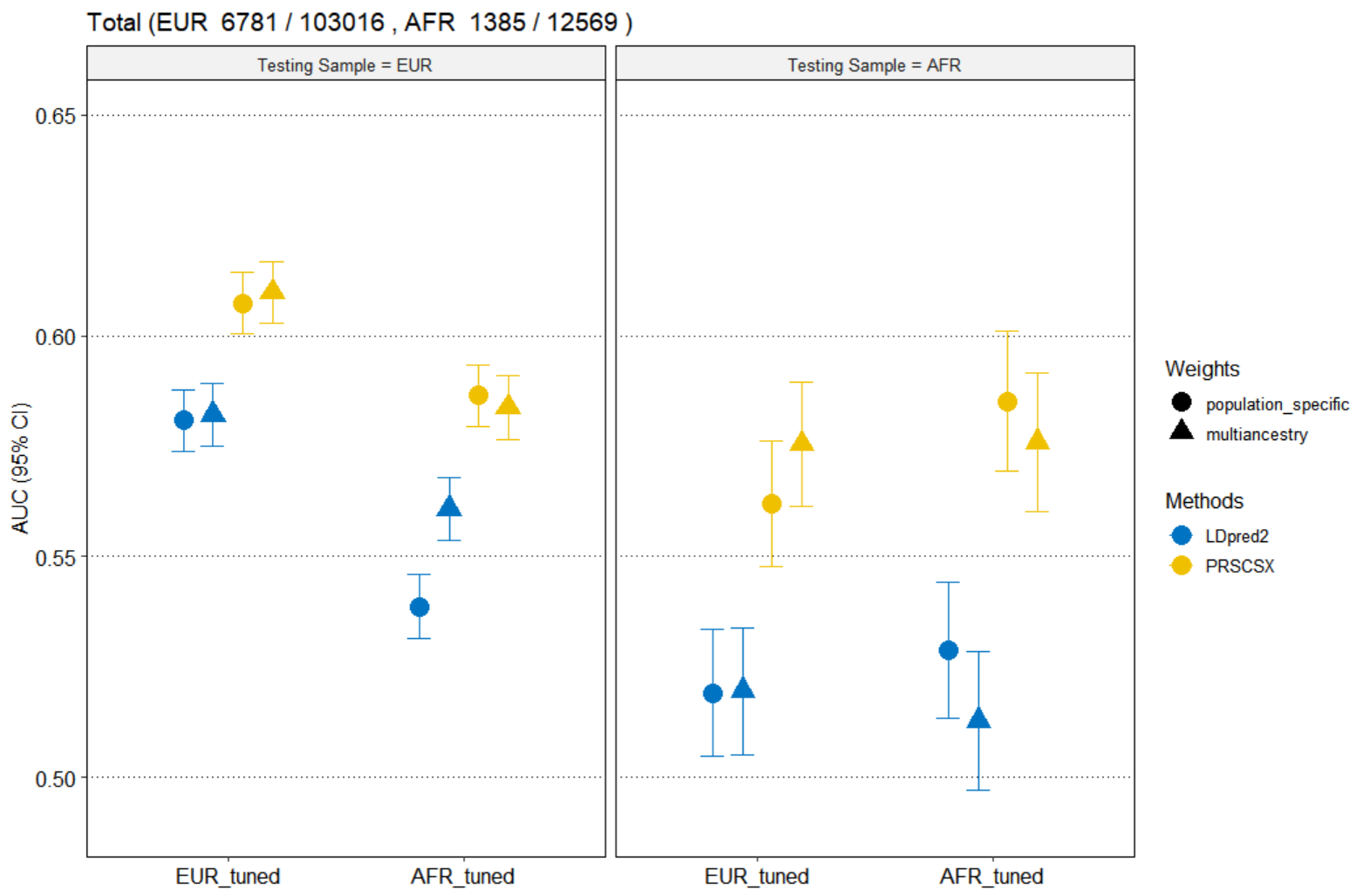
AFR: MGI (EUR_tuned), PMBB, UCLA
